# Examining the causal genetic effects of substance use on aging

**DOI:** 10.1101/2024.10.14.24315472

**Authors:** Jared V. Balbona, Paul Jeffries, Aaron J. Gorelik, Elliot C. Nelson, Ryan Bogdan, Arpana Agrawal, Emma C. Johnson

## Abstract

Substance use shortens lifespan, impedes health, and accelerates the biological aging process. We found widespread genetic correlations between alcohol, tobacco, cannabis, and opioid use and use disorders with indices of aging across the lifespan. There was evidence of tobacco and alcohol use and use disorders causally impacting physical, cognitive, and biological aging, with the effects of alcohol being more dependent on quantity of consumption; evidence of reverse causality was scant.

## Main Text

Chronic substance use and substance use disorders (SUDs)—psychiatric conditions marked by interpersonal, physiological, and psychological problems resulting from the loss of control over substance use—are among the leading causes of preventable death globally, placing a substantial burden on both individuals and society^1^. Despite this, not all individuals who use substances or have SUDs suffer premature mortality as a result. While many experience recovery and remission, others continue using substances maladaptively into later life, as evidenced by rising SUD rates among older adults^2^. As global life expectancy and substance availability have increased^2^, it is crucial to understand if and how substance involvement directly complicates biological aging and health in later life.

Psychoactive substances can directly and indirectly impact physical health, with evidence linking prolonged substance exposure to discrepancies between individuals’ chronological and biologically estimated ages^3^. What is less well-understood, however, is the extent to which substance involvement *causally* modifies physical and biological aging (e.g., via neurotoxic effects) as opposed to both SUDs and accelerated aging resulting from shared predisposing factors (e.g., neighborhood disadvantage, medical stigma). Given the heritable nature of substance use and most aging metrics, genetically-informed study designs are well-suited to estimate the extent of shared genetic vulnerability to both domains and, after accounting for horizontal pleiotropy, to evaluate support for causal inference^4^.

In the present study, we assessed whether substance involvement (i.e., use and use disorders) is genetically correlated with physical, cognitive, and biological aging, and whether such associations reflect shared predisposition and/or causal influences of substance involvement. To this end, we calculated genetic correlations between 8 substance use and SUD phenotypes (drinks per week, problematic alcohol use, smoking initiation, tobacco use disorder, cannabis ever use, cannabis use disorder, opioid use disorder, and a multivariate SUD factor (mvSUD)) and 8 aging traits (Alzheimer Disease, frailty index, healthspan, parental lifespan, GrimAge, leukocyte telomere length, brain-age gap, and a multivariate aging factor (mvAge)) using Linkage Disequilibrium Score Regression^5^ (LDSC; **Online Methods)**. We then employed a series of Mendelian Randomization (MR) based approaches^6^, which utilize genetic variants as instrumental variables in order to estimate the causal effect of an “exposure” (e.g., SUD) on an “outcome” (e.g., aging; **Online Methods**). Briefly, MR leverages the random assortment of genes from parents to offspring, thereby ensuring that confounders are randomly distributed across the “treatment” and “control” groups (those who do and do not have a copy of the effect allele, respectively) in a manner analogous to a randomized control trial. By reducing the potential for confounds in this way, MR-based approaches can disaggregate horizontal pleiotropy (the influence of a genetic locus on two separate, independent traits) from vertical pleiotropy (the direct causal effect of an exposure on an outcome; *Fig. 1*), allowing researchers to infer causality in situations where pleiotropy is expected and randomized control trials are not feasible.

**Figure 1.**
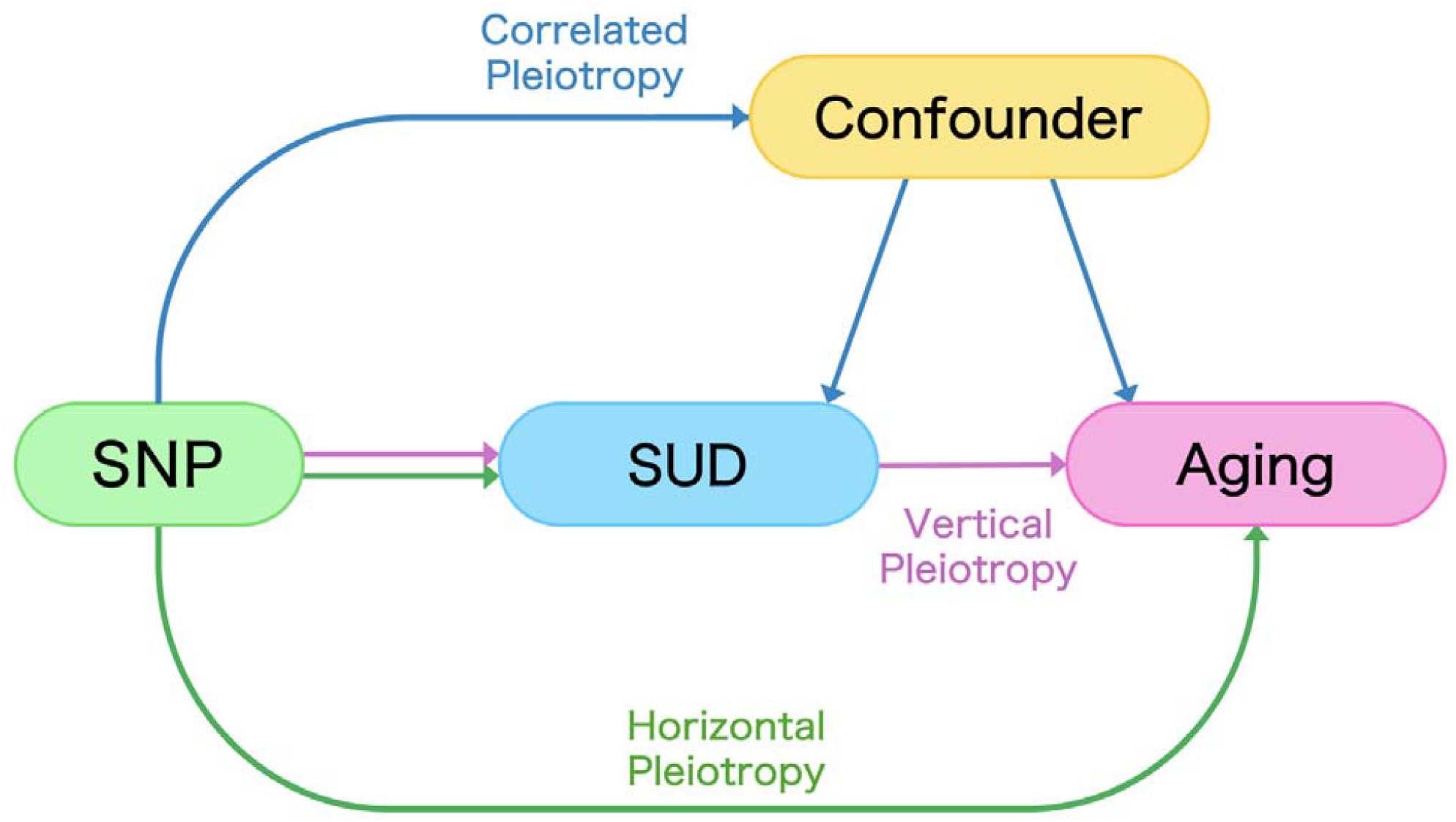
Vertical pleiotropy occurs when a single nucleotide polymorphism (SNP) influences multiple traits through a direct causal pathway, where the SNP affects one trait that in turn affects another. In contrast, horizontal pleiotropy happens when a single SNP independently influences multiple traits through separate and distinct mechanisms, without one trait affecting the other. Correlated pleiotropy involves a situation where a single gene affects multiple traits that are correlated, often because the traits share common genetic or environmental influences.

## Genetic Correlations

LDSC revealed widespread correlations between substance involvement and aging phenotypes, with genetic correlations ranging from *r*_*g*_ = −.50 (tobacco use disorder and parental lifespan) to *r*_*g*_ = .55 (smoking initiation and GrimAge), with a median absolute genetic correlation (|*med _rg_*|) of .18 across all between-category trait pairs (*Fig. 2*). mvSUD was significantly associated with all aging indices (|*med _rg_* | = .36) and mvAge was significantly associated with all substance involvement phenotypes, except cannabis ever use (|*med _rg_* | = .35). Similarly, parental lifespan was significantly correlated with all substance use measures (|*med _rg_* | = .36), while problematic alcohol use (|*med _rg_* | = .27), smoking initiation (|*med _rg_* | = .36), and tobacco use disorder (|*med _rg_* | = .36) were each associated with 7 of 8 aging metrics. In contrast, despite cannabis use disorder showing moderate-to-high associations with most aging measures (|*med _rg_* | = .33), cannabis ever use was only modestly correlated with parental lifespan (r g = .10) and no other aging index.

**Figure 2.**
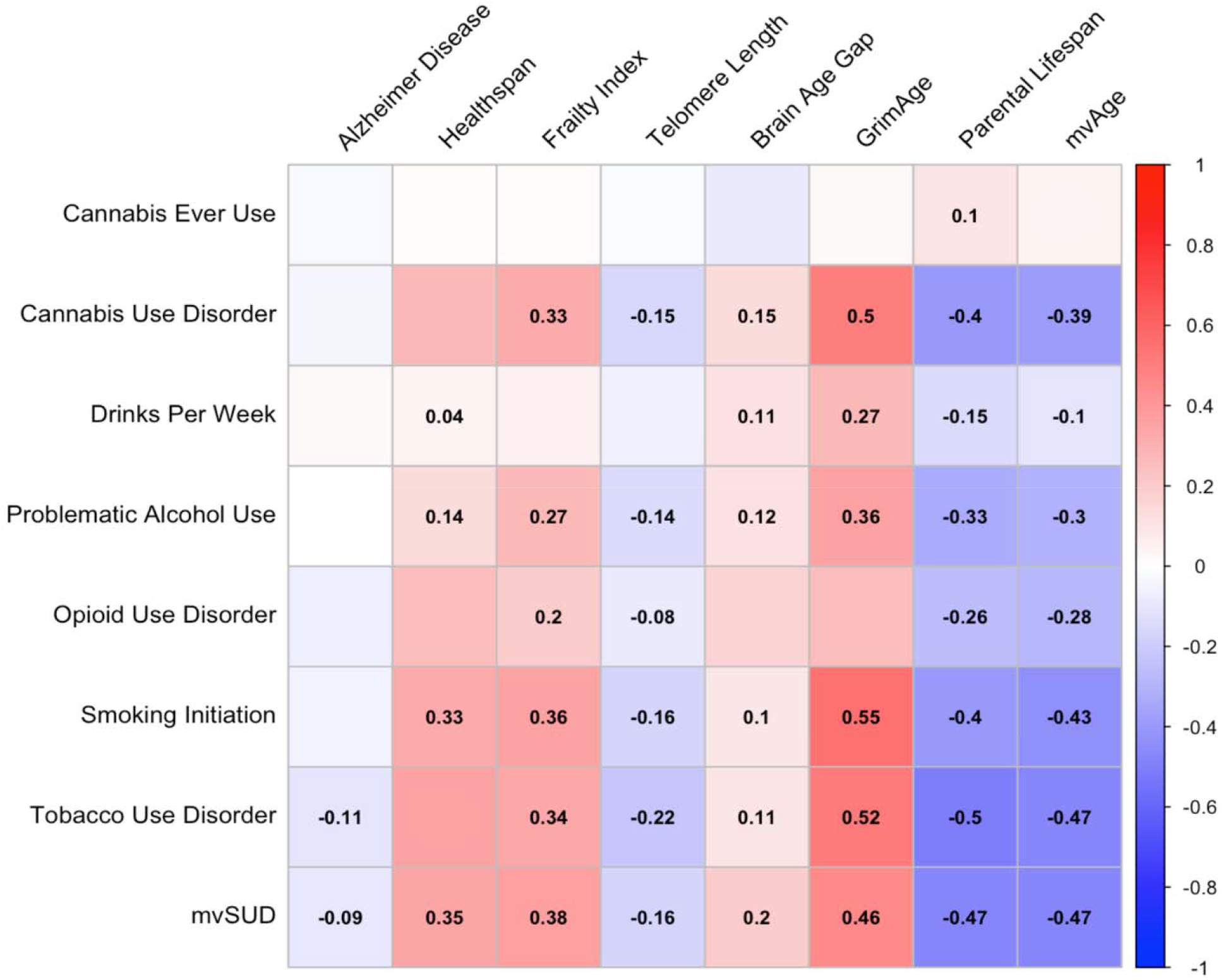
Genome-wide genetic correlations between trait pairs, calculated using LDSC. Coefficients are provided for correlations reaching FDR-adjusted significance (all correlations are available in Supp Table 2). As shown, correlations were widespread, particularly for smoking and alcohol phenotypes, parental lifespan, mvSUD. and mvAge.

## Mendelian randomization

For each of the significantly genetically correlated trait pairs, we evaluated evidence for genetic causation using Causal Analysis Using Summary Effect Estimates^7^ (CAUSE) and Mendelian Randomization Pleiotropy RESidual Sum and Outlier^8^ (MR-PRESSO), with the two approaches showing consistent agreement with one another despite their differing assumptions (*Fig. 3*). MR-PRESSO—which detects and removes potential pleiotropic outlier SNPs—found significant causal effects for smoking initiation and tobacco use disorder on 6 and 7 aging-related indices, respectively; problematic alcohol use and drinks per week, meanwhile, both showed causal effects on 3 aging metrics. All of these effects persisted after outlier SNPs were removed, indicating that they are likely due to vertical pleiotropy rather than outlier SNP-induced horizontal pleiotropy. Similarly, CAUSE results provided evidence that genetic liability for problematic alcohol use, smoking initiation, tobacco use disorder, and mvSUD may be causal for several aging outcomes, including frailty, GrimAge, and parental lifespan (*Supp Table 4)*; across these traits, CAUSE found that a causal effect model fit the data significantly better than did a shared effects model (*Supp. Table 5)*. Finally, we examined the effects of aging on substance use using both MR-PRESSO and CAUSE, finding little evidence of reverse causality from either approach.

**Figure 3.**
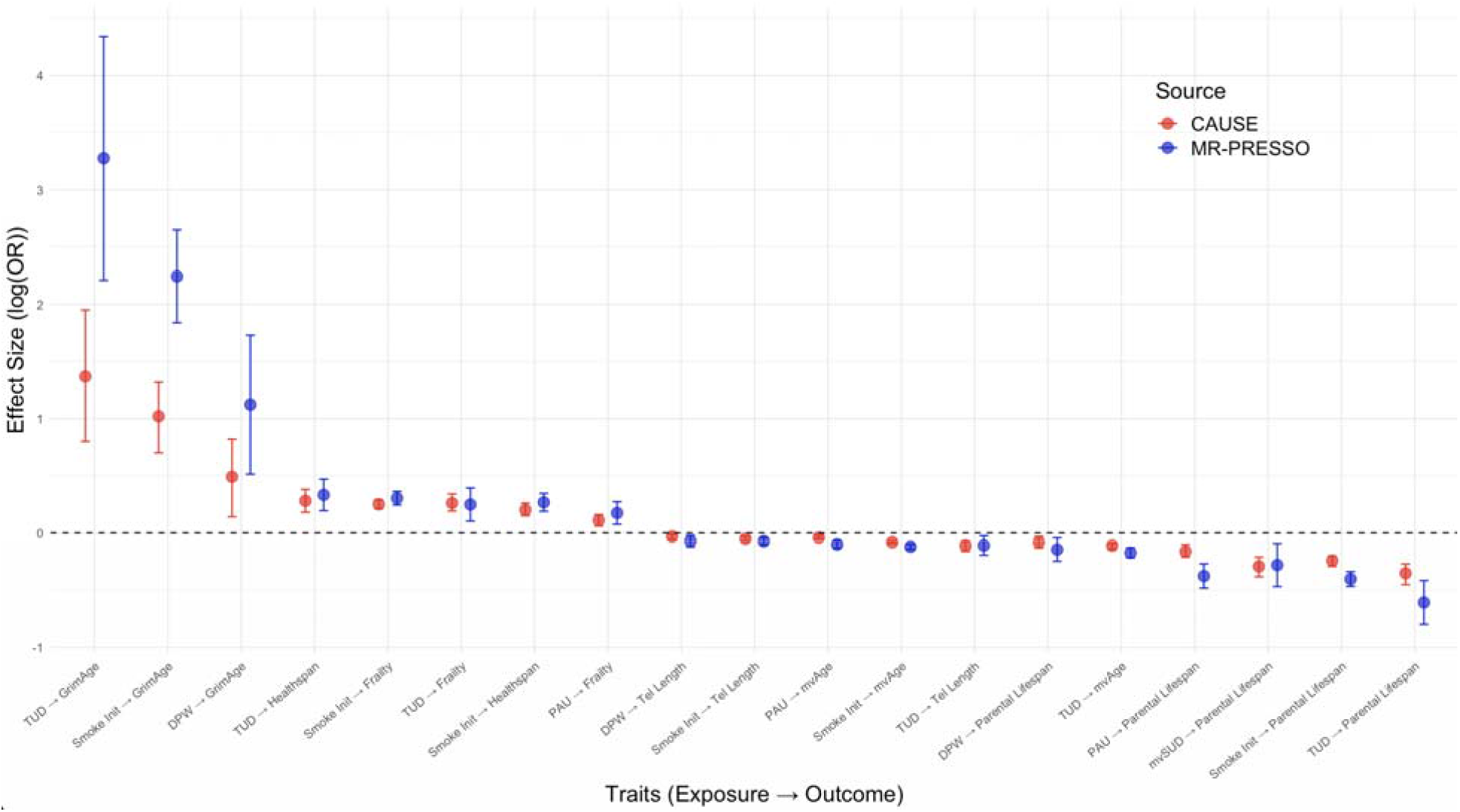
CAUSE and MR-PRESSO effect size estimates (log(OR)) for significant trait pairs, with significance being based upon an FDR-corrected *p*-value threshold. Results from both approaches are largely in agreement with one another, with the error bars (representing 95% confidence intervals) overlapping for nearly all trait pairs.

Substance use and SUDs contribute to a majority of the leading causes of death and disability across the world (e.g., chronic obstructive pulmonary disease, cardiovascular disease, cancer)^9^. We found widespread evidence of genetic correlations between substance use/ SUDs and aging metrics, with MR results suggesting that both tobacco and alcohol use are causally implicated in shortening lifespan, deterring geriatric health, and modifying markers of biological youth. Notably, despite the causal effects of smoking initiation largely mirroring those of tobacco use disorder, we found that problematic alcohol use had a more significant impact on lifespan and mvAge than did drinks per week. These findings suggest that the effects of alcohol on aging may be quantity dependent, and thus highlight the importance of parsing the effects of quantitative substance exposure from disordered use to more fully understand their multifaceted impact on health. Furthermore, our scant evidence for reverse causality (i.e., aging impacting substance use) suggests that the reported increase^2^ in SUD rates among older adults are not necessarily occurring in response to the physical and mental challenges of aging, as has been previously theorized^10^.

While our null MR findings (e.g., for cannabis, opioids, and some aging metrics) suggest that the observed genetic correlations between these SUD exposures and aging outcomes represent either common genetic mechanisms or third variable confounding, some caveats remain noteworthy. First, we cannot exclude the possibility that some null findings—particularly those involving cannabis use and Alzheimer Disease—were due in part to smaller GWAS sample sizes and a consequent lack of power to detect effects. Relatedly, we cannot dismiss the potential consequences of lower rates of recent SUDs in older individuals with health conditions (the “sick quitter” phenomenon^11^), survivorship/ selection bias, unmodeled interactive effects, or non-representative GWAS samples. Additionally, all aging-related summary statistics were derived from GWAS that only included individuals of European genetic ancestry, limiting the generalizability of these findings to other ancestries (**Online Methods**).

Stigma associated with the acknowledgement of SUDs and the consequent reluctance to query older adults about their substance use hinder effective and tailored interventions in this age group^12^. Our analyses suggest that prevention of substance use may reduce markers of biological aging and promote geriatric well-being and longevity. Additionally, these results underscore the importance of integrating genetic insights into public health strategies to effectively reduce the burden of SUDs across the lifespan.

## ONLINE METHODS

### Data Sources and Phenotypes

Genetic association estimates were derived using publicly available summary statistics based on individuals of European ancestry (see *Supp. Table 1 for full descriptions for each trait/ cohort)*. In total, eight substance use traits (problematic alcohol use, drinks per week, smoking initiation, cannabis ever use, cannabis use disorder, opioid use disorder, tobacco use disorder, and a multivariate substance use factor (mvSUD)) and eight aging traits (telomere length, Alzheimer Disease, healthspan, brain age gap, parental lifespan, frailty index, GrimAge, and a multivariate aging factor (mvAge)) were analyzed.

### Study Design

All pairwise combinations of substance use and aging traits were first analyzed using LD score regression. Subsequently, causality between all significantly correlated pairs of traits were analyzed using both MR-PRESSO and CAUSE.

### Genetic Instrument Selection and QC

All included SNPs were non-palindromic, autosomal, and genome-wide significant (*p* ≤ 5×10^−8^). To exclude variants adjacent to the variants of interest, SNPs were clumped based on their linkage disequilibrium (*r*^2^ = .01; *p* < 5×10^−3^) within a genomic window of 10,000 kb, with the 1000 Genomes European LD estimates serving as our reference panel. Additionally, each non-effect allele was aligned to the human genome reference sequence (build 37) to ensure that the effect of a given SNP is corresponding to the same allele for both the exposure and the outcome.

Finally, the MR-PRESSO outlier test was used to exclude any potential outlier variants from the MR-PRESSO analyses.

### Linkage Disequilibrium Score (LDSC) Regression

LDSC regression was utilized to examine the genetic correlations between all pairs of traits, both within-category (i.e., aging-aging, SUD-SUD) and between-category (SUD-aging). Genetic correlation estimates from LDSC regression are not biased by sample overlap, but rather are able to estimate and account for the degree of overlap; as such, it is able to theoretically estimate the correlation between traits that is due to similarities in genetic architecture, rather than sampling artifacts. Because several of our traits are non-independent of one another (e.g., problematic alcohol use and drinks per week), an FDR-corrected *p*-value was employed to reduce the risk of type I errors.

### Mendelian Randomization Methods

A primary assumption of traditional MR-based approaches is that the genetic instruments should influence the outcome exclusively through the exposure of interest (i.e., the instrument exclusion restriction). However, this assumption is commonly violated due to the presence of horizontal pleiotropy, both when that the pleiotropy is correlated (i.e., when traits share a genetically influenced causal pathway) and when it is uncorrelated (when the genetic factor influences the exposure and outcome via distinct mechanisms). Therefore, the present study implements a two-pronged MR approach to assessing the causal impact of substance use on aging.

First, MR-PRESSO was conducted using the MRPRESSO R package version 1.0 (https://github.com/rondolab/MR-PRESSO). Here, we present findings from the MR-PRESSO raw tests, except in cases in which the MR-PRESSO distortion test (indicating a significant influence of outlier variants) is FDR significant; in such cases, the MR-PRESSO outlier-corrected values are given instead. Additionally, we performed CAUSE using the R package cause version 1.2.0 (https://github.com/jean997/cause). CAUSE assumes that genetic relationships between traits are a mixture of both vertical and correlated pleiotropy, and thus evaluates the relative contributions of both. MR-PRESSO, meanwhile, detects and removes SNPs with evidence of horizontal pleiotropy, thus minimizing bias in its estimate of SNP-based vertical pleiotropy.

## Data Availability

All data sources in the present work are contained in the manuscript

https://github.com/JaredBalbona/sud_aging_pleiotropy.git

## Notes

### Competing Interest Statement

The authors have declared no competing interest.

### Funding Statement

This study was funded by the following grants: R01DA054869, T32DA007261, K01DA051759.

### Author Declarations

The present study utilized publicly available genome wide association summary statistics. The data sources for each are available in our supplementary materials

